# Self-testing for the detection of SARS-CoV-2 infection with rapid antigen tests

**DOI:** 10.1101/2021.02.21.21252153

**Authors:** J. J.J.M. Stohr, V. F. Zwart, G. Goderski, A. Meijer, C. R.S. Nagel-Imming, M.F.Q. Kluytmans-van den Bergh, S. D. Pas, F. van den Oetelaar, M. Hellwich, K. H. Gan, A. Rietveld, J.J. Verweij, J. L. Murk, W. van den Bijllaardt, J. A. J. W. Kluytmans

## Abstract

**Introduction:** Self-testing for COVID-19 infection with lateral flow assay SARS-CoV-2 rapid antigen detection tests (RDT), provides rapid results and could enable frequent and extensive testing in the community, thereby improving the control of SARS-CoV-2. The objective of this study is to evaluate the performance of self-testing using RDT without assistance.

**Methods:** Participants visiting a municipal SARS-CoV-2 testing centre, received self-testing kits containing either the BD Veritor System (BD RDT) or Roche SARS-CoV-2 antigen detection test (Roche RDT). Oro-nasopharyngeal swabs were collected from the participants for qRT-PCR testing. As a proxy for contagiousness, viral culture was performed on a selection of qRT-PCR positive samples to determine the Ct-value at which the chance of a positive culture was dropping below 0.5 (Ct-value cut-off). Sensitivity and specificity of self-testing were compared to qRT-PCR with a Ct-value below the Ct value cut-off. Determinants independently associated with a false-negative self-test result were determined.

**Results:** A total of 3,215 participants were included (BD RDT n=1604; Roche RDT n=1611). Sensitivity and specificity of self-testing compared to the qRT-PCR results with Ct-value below the Ct-value cut-off was 78.0% (95% CI:72.5-82.8) and 99.4% (95%CI: 99.0-99.6) respectively. Determinants independently associated with a false-negative self-testing results were: higher age, low viral load and finding self-testing difficult.

**Discussion:** Self-testing using currently available RDT’s has a high specificity and relatively high sensitivity to identify individuals with a high probability of contagiousness. The performance of two tests were comparable. This application has the potential for frequent and extensive testing which may be an aid to lift restrictions to society while controlling the spread of SARS-CoV-2.

## Introduction

In December 2019, severe acute respiratory syndrome coronavirus type 2 (SARS-CoV-2) was first identified as a causal agent of viral pneumonia in Wuhan, China (1,2). Since then, SARS-CoV-2 has spread across the globe causing millions of cases of coronavirus disease 2019 (COVID-19). Extensive testing of individuals who are potentially infected with SARS-CoV-2, has a central role in efforts to mitigate the spread of SARS-CoV-2 (3,4). Current testing strategies for SARS-CoV-2 infection are hampered by the necessity to deploy qualified personnel to collect the sample, perform the tests and to interpret the test results. Self-testing for SARS-CoV-2 infection, where patients collect the sample, perform the test and interpret the result themselves, could enable massive testing in the community, thereby improving the control of SARS-CoV-2 (5,6). Due to their ease-of-use, short turn-around time, and low-costs, lateral flow assay SARS-CoV-2 antigen tests (RDT), could be suitable candidates for self-testing for SARS-CoV-2 infection (7–9). Studies on the performance of these antigen rapid detection tests have shown promising results when samples were collected and performed by qualified personnel (7–9). Recent reports have shown comparable performance results between SARS-CoV-2 antigen rapid tests performed on nasal mid-turbinate samples and nasopharyngeal samples, and have established the achievability of nasal mid-turbinate self-sampling under supervision (10–12). At present, data on the performance of self-testing with SARS-CoV-2 antigen rapid detection tests (RDT) is limited to comparisons with real-time reverse transcription polymerase chain reaction (qRT-PCR) detection of SARS-CoV-2 RNA. qRT-PCR detects intact virus but also non-transmittable SARS-CoV-2 RNA (14,15), and could, therefore, overestimate the number of contagious patients. Other reports have tried to overcome this limitation by stratifying the results for the cycle threshold value (Ct-value) of the RT-PCR (7,8). However, the Ct-value at which patients are expected to be no longer contagious is not known for most qRT-PCR assays and patient populations. The objective of this study is to evaluate the performance of self-testing using two commercially available lateral flow SARS-CoV-2 antigen assays i.e., BD Veritor System for Rapid Detection of SARS-CoV-2 (Becton Dickinson company, USA) (BD RDT) and Roche SARS-CoV-2 antigen detection test (Roche, Switzerland) (Roche RDT), for the detection of contagious COVID-19 patients in the community.

## Methods

### Study design and participants

This manufacturer-independent cross-sectional study was conducted from December 23, 2020, to January, 17, 2021, in the test centre of the Municipal Health Services in Tilburg, Noord-Brabant, the Netherlands. In the Netherlands community testing for SARS-CoV-2 is coordinated by the MHS. Adults above the aged 18 years or older who presented at the test centre, were able to understand the written instructions in Dutch and provided verbal informed consent procedure and contact information (e-mail address and telephone number) were deemed for inclusion.

### Study procedure

Eligible participants were randomly allocated to either a test lane distributing the BD RDT self-testing kit or a test lane distributing the Roche RDT self-testing kit. Participants received a small bag with the self-testing kit, were instructed to perform the SARS-CoV-2 self-test immediately after arrival at-home and were asked to provide their e-mail address and telephone number. At the test centre, the standard method for SARS-CoV-2 testing was carried out. Oro- and nasopharyngeal swabs were collected from the participants by a trained member of the MHS and suspended in 3mL gelatin-lactalbumin-yeast virus transport (GLY) medium (Mediaproducts, the Netherlands). The suspended swabs were sent to Microvida Laboratory for Medical Microbiology and Immunology, Tilburg, the Netherlands, for qRT-PCR testing within 4 hours after sample collection. Participants received an e-mail with to a survey. When the participant did not complete the survey within two hours following inclusion, the participant was telephoned by a member of the research team to fill in the survey form jointly. If the participant did not yet perform the self-test when being telephoned, the member of the research team asked the participant to perform the test and fill in the survey form sent via e-mail. Participants were not assisted during the self-testing procedure.

### Self-testing

Participants received a self-testing package containing either a BD RDT or a Roche RDT, a flocked swab, a foldable cardboard test frame, and a written and illustrated booklet including general information on the study and an instruction on how to collect a mid-turbinate nasal sample, how to perform the test and how to interpret the test result. This instruction included a QR-code link to a two-minute online video illustrating mid turbinate self-sampling and self-testing using the BD RDT (http://www.corona-test-instructies.nl/) and Roche RDT (http://www.coronatest-instructies.nl/) (**Supplementary methods S1** and **Supplementary methods S2**).

The BD RDT and Roche RDT test are both chromatographic lateral flow immunoassays for the qualitative detection of SARS-CoV-2 nucleocapsid antigen in swabs from individuals who are suspected of COVID-19 within the first 5 days of symptom onset. Both tests are currently not approved or validated for self-testing purposes.

### Survey

Participants were asked to provide their age, gender, highest level of education, information regarding current and recent COVID-19 related symptoms, and respond to statements regarding the self-test procedure (**Supplementary method S3**).

### qRT-PCR

Nucleic acids extraction and real-time RT-PCR for SARS-CoV-2 were performed on combined oro- and nasopharyngeal swabs suspended in 3mL GLY-medium either with the CE-IVD labelled ‘Alinity M SARS-CoV-2 Assay’ (Abbott) (AA) with a combined N-gene and RdRP-gene target, according to the manufacturer’s instructions or with an internally controlled, E-gene target, lab-developed assay (LDA) using the QIAsymphony Sample Processing and Rotorgene amplification system (Qiagen, Hilden, Germany), as described previously by Sikkema et al. (16).

### Viral culture

Samples with a positive qRT-PCR result, that were collected before January 12, 2021, were frozen at −80° C within 24 hours after sample collection and transported on dry-ice to the laboratory of the Dutch National Institute for Public Health and the Environment (RIVM) for viral culture. All virus isolation work was performed at Biosafety Level-3. After thawing the RT-PCR positive samples at room temperature the tubes were vortexed for 1 minute at 1000 RPM to homogenise the sample. Subsequently the samples were centrifuged for 10 minutes at 7500 RPM to pellet fungi or bacteria, minimising the risk of contamination. From each sample 250 µl supernatant was inoculated on 24-wells plate with 3 days old VERO-E6 (ATCC CRL-1586) monolayer in 1 ml Dulbecco’s Modified Eagle Medium with 5% fetal bovine serum, 100 units penicillin and streptomycin/ml and 100 units nystatin/ml (DMEM) and incubated at 34 °C and 5% CO2 for 7 days. After 7 days presence of cytopathic effect (CPE) was recorded and a next passage was done on 24-wells with VERO-E6 cells monolayer with 1 ml DMEM by transferring 25 µl supernatant for wells with CPE and 250 µl for wells with no CPE and for wells contaminated with bacteria/fungi after the supernatant was filtered through a 0.22 µm filter. After incubating for 7 days at 34 °C and 5% CO2 CPE was recorded and to confirm the culture is positive a qRT-PCR was performed. From each culture 200 µl was mixed with 275 µl Roche MagNAPure96 (MP96) lysis buffer which includes 25 µl Equine Arteritis Virus (EAV) internal control and 450 µl was extracted on a MP96 Instrument (Hoffmann-La Roche, Basel, Switzerland) using the MP96 DNA and Viral NA Small Volume Kit and eluted in a volume of 50 µl. The E-gene/EAV multiplex qRT-PCR was used to test inhibition of EAV amplification and to confirm the culture was positive in the E-gene qRT-PCR. E-gene primers and probes were as described by Corman et al. (17) and EAV as described by Scheltinga et al. (18). All tests are run on the Light Cycler 480 I using TaqMan® Fast Virus 1-Step Master Mix (ThermoFisher Scientific) in a temperature cycling programme at 60° annealing/elongation according to the manufacturer’s instructions.

### Sample size

At the start of the study, the diagnostic accuracy of self-performed rapid antigen tests was unknown. We assumed the diagnostic accuracy to be lower than when performed by professionals, and based the sample size calculation on an expected sensitivity of 80% for infectious individuals, with a margin of error of 7%, type I error of 5% and power of 90%. Hence, the minimum number of participants with a positive qRT-PCR test was 140 per LFA arm. We will monitor the qRT-PCR test positivity percentage over time and adjust recruitment if needed.

### Statistical analysis

The primary outcome was the sensitivity and specificity of the RDT compared with qRT-PCR with a Ct-value lower than or equal to a Ct-value cut-off that correlated with at least a 50% chance of recovering a viable virus using viral culture. Therefore, an univariate logistic regression analyses was performed to determine the Ct-value at which the chance (p) of having a positive viral culture was p = 0.5 (Ct-value cut-off) for both the AA and LDA qRT-PCR with Statsmodels v0.12.2. This Ct-value cut-off was used as a proxy for a contagious SARS-CoV-2 viral load. Univariate and multivariate logistic regression analyses were performed using Statsmodels v0.12.2 to examine whether following variables were independently associated with a false negative result in self-test for COVID-19 as compared to the qRT-PCR: Antigen rapid test used (BD RDT or Roche RDT), Age, Sex, current COVID-19 related symptom, Ct-value lower than the Ct-value at which the chance (p) of having a positive viral culture was p = 0.5, and the responses to the statements included in the survey. Responses to the survey statements were redefined as dichotomous variables (responses 1 (totally agree) and 2 (agree) as 1 and responses 3 (disagree) and 4 (totally disagree) as 0). Variables were included in the multivariate analyses when p < 0.2 in the univariate analyses. Additionally, for the samples with a viral culture result, a composite reference standard for expected contagious COVID-19 infection was created. This composite reference standard was defined as positive when having a positive result in at least 2 out of the following 3 tests: Viral culture, qRT-PCR, and Antigen rapid test. Adjusted Wald confidence intervals (CI) for the sensitivity and specificity of self-testing as compared to RT-PCR overall, RT-PCR with an expected contagious viral load, and composite reference standard were calculated using Scipy v.1.17.0. Inconclusive results in the qRT-PCR were excluded from all further analysis. Samples of participants with an inconclusive result in the self-test were excluded when determining the sensitivity and specificity of self-testing, were included for determining the Ct-value cut-off, and interpreted as not false negative when determining the variables associated with a false negative result.

### Ethics

The study protocol was reviewed by the Dutch ‘Medical research Ethics Committees United’ (MEC-U). The study was judged to be beyond the scope of the Dutch medical scientific research act (WMO). A waiver of written informed consent was granted to enable the required high flow of individuals in the test centre and prevent any safety hazard associated with the handling of documents obtained from possibly infectious participants.

## Results

In total, 3,529 eligible participants were included in the study, of whom 3,215 (91.1%) were willing and able to complete and share the survey results (**Figure 1**). Of the 3,215 respondents, 1,604 (49.9%) received a self-testing package containing the BD RDT and 1,611 (50.1%) the self-testing package containing the Roche RDT (**Figure 1**). No difference in baseline characteristics of the participants who completed and shared their survey result were detected between the two groups of respondents (**Table 1**). Because of sample loss the RT-PCR was not done in 11 (0.3%) out of the 3,215 samples. The AA and LDA were performed in 994 (30.9%) and 2210 (68.7%) samples respectively. Results of the qRT-PCR were positive in 377 (11.7%) samples, negative in 2,824 (87.8%), and inconclusive in 3 (0.1%). Out of the 377 samples with a positive qRT-PCR result, 289 were sent for viral culture (AA: n=85 samples; LDA: n=204 samples). Of those, 178 (61.6%) had a positive culture result (**Supplementary table S1**). For both the LDA and the AA the Ct-values in the samples with a positive viral culture (median Ct-value AA: 17.9 (Interquartile range (IQR)1-IQR3: 16.1-19.8) and median Ct-value LDA: 18.1 (IQR1-IQR3: 16.4-20.5)) were lower than in the samples with a negative viral culture (median Ct-value AA: 28.3 (IQR1-IQR3: 25.1-33.1) and median Ct-value LDA: 31.0 (IQR1-IQR3: 25.0-34.8)) (**Figure 2**). Based on the univariable logistic regression model the Ct-value cut-off where the chance for a positive viral culture was smaller than 0.5 was 23.0 (95%CI: 16.0-43.0) for the AA (pseudo-R^2^: 0.638) and 24.5 (95%CI: 20.4-33.6) for the LDA (pseudo-R^2^: 0.570) (**Supplementary figure S1, Supplementary figure S2**).

**Figure 1.**
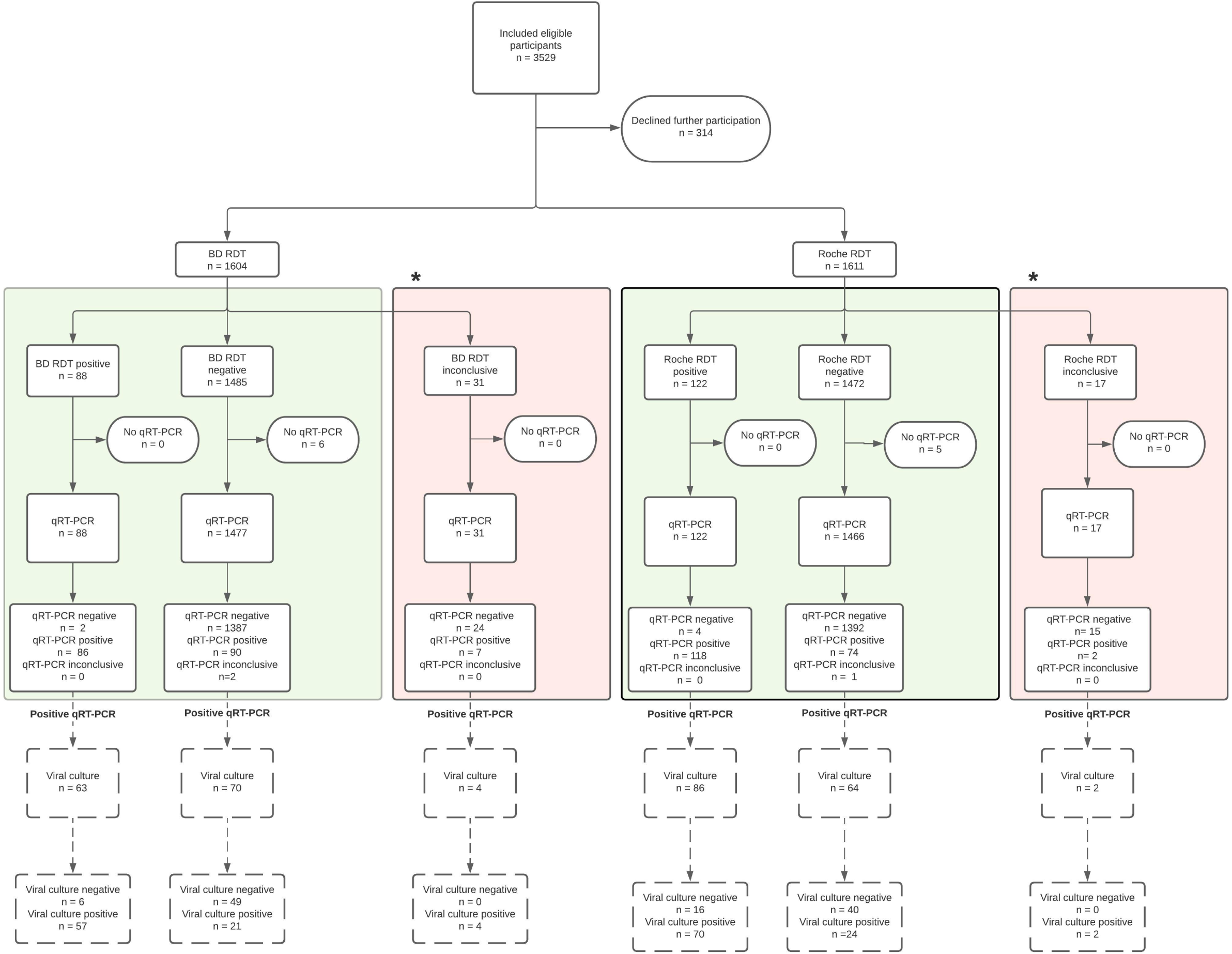
Participant flow diagram. *Samples of participants were included in the analysis determining the Ct-value cut-off where the p for a positive viral culture was smaller than 0.5 but not in the analysis determining the sensitivity and specificity of self-testing.

**Table 1.** Baseline characteristics study participants; IQR: interquartile range.

|  | Overall | Test type |  |
| --- | --- | --- | --- |
|  |  | BD RDT | Roche RDT |
|  | n = 3215 | n = 1604 | n = 1611 |
| Age, median (Years) (IQR-IQR) | 41 (29-54) | 41 (28-54) | 41 (29-54) |
| Gender, n (%) |  |  |  |
| male | 1409 (43.8) | 720 (44.9) | 689 (42.8) |
| female | 1806 (56.2) | 884 (55.1) | 922 (57.2) |
| Highest level of education, n (%) |  |  |  |
| Elementary school | 63 (2.0) | 35 (2.2) | 28 (1.7) |
| Highschool | 1618 (50.3) | 835 (52.1) | 783 (48.6) |
| Bachelor degree | 1075 (33.4) | 516 (32.2) | 559 (34.7) |
| Masters degree or higher | 459 (14.3) | 218 (13.6) | 241 (15.0) |
| Currently symptoms COVID-19 infection, n (%) |  |  |  |
| Yes | 2226 (69.2) | 1119 (69.8) | 1107 (68.7) |
| No | 989 (30.8) | 485 (30.2) | 504 (31.3) |
| Symptoms COVID-19 infection in past 3 weeks, n (%) |  |  |  |
| Yes | 201 (6.3) | 99 (6.2) | 102 (6.3) |
| No | 3014 (93.7) | 1505 (93.8) | 1509 (93.7) |
| No symptoms COVID-19 infection, n (%) |  |  |  |
| Yes | 788 (24.5) | 386 (24.1) | 402 (25.0) |
| No | 2427 (75.5) | 1218 (75.9) | 1209 (75.0) |

**Figure 2.**
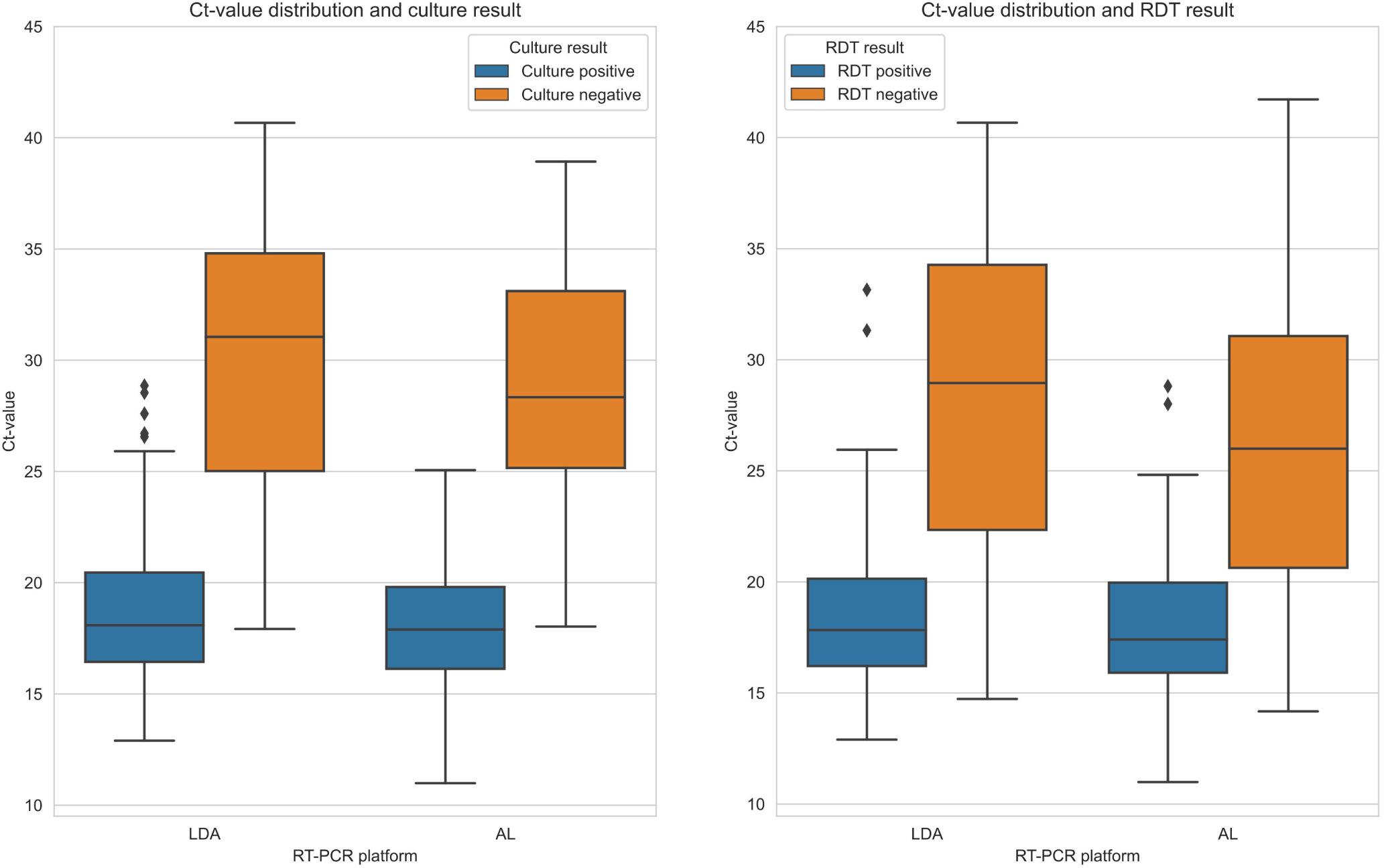
Ct-value distribution per qRT-PCR platform for viral culture positive and viral culture negative samples and for RDT positive and RDT negative samples.

Out of the total of 3,215 RDT self-tests performed, 210 (6.5%) tests were positive (BD RDT: n=88; Roche RDT n=122), 2,957 (92.0%) tests were negative (BD RDT: n=1485; Roche RDT n=1472), and 48 (1.5%) tests yielded an inconclusive result (BD RDT: n=31; Roche RDT n=17) (**Figure 1**;). No difference in responses to the survey statements between participants receiving a BD or Roche RDT was detected (**Supplementary table S2**). The sensitivity and specificity of self-testing compared to qRT-PCR were 55.4% (95%CI: 50.3-60.4) and 99.8% (95%CI: 99.5-99.9) respectively (Table 2; Supplementary table S3). For both the Roche RDT and BD RDT the specificity was high, being 99.7% (95%CI: 99.3-99.9) and 99.9 % (95%CI: 99.5-100) respectively. The sensitivity of self-testing using the Roche RDT (61.5% (95%CI: 54.4-68.1)) was higher than when using the BD RDT (48.9% (95%CI: 41.6-56.2)) (**Table 2; Supplementary table S3**). When only samples with a Ct-value in the qRT-PCR below the previously defined Ct-value cut-off were considered positive, the sensitivity of self-testing using both the Roche RDT and BD RDT increased to respectively 80.1% (95%CI: 72.7-86.0) and 75.5% (95%CI: 66.6-82.6), with a relatively small decrease in specificity (BD RDT: 99.7% (95%CI: 99.2-99.9); Roche RDT: 99.1% (95%CI: 98.5-99.5)) (**Table 2; Supplementary table S3**). When comparing the result of the self-test to the composite reference standard, the sensitivity was 76.8% (95%CI: 70.4-82.2) (**Table 2; Supplementary table S4**). This sensitivity was similar to that comparing self-testing results to PCR positive samples with a Ct-value below the Ct-value cut-off. The association between a false negative test result in the self-test as compared to the qRT-PCR was investigated for 14 variables using a univariable logistic regression model (**Table 3**). Out of these 14 variables, 7 variables were associated with the occurrence of a false negative self-test result with a p value < 0.2 and were included in the multivariate analysis (**Table 3**). A higher age, judging self-testing as difficult and a Ct-value in the RT-PCR higher than the cut-off value were independently associated with the occurrence of a false negative self-test results (**Table 3**. In participants with an age below the median age (41), that found self-testing very easy (10) and with a Ct-value in the RT-PCR lower than the cut-off value, the sensitivity of self-testing increased to 85.0% (95%CI: 70.5-93.3).

**Table 2.** Sensitivity and specificity of the self-testing compared to qRT-PCR, qRT-PCR with a Ct-value smaller than the cut-off and to the composite reference standard. Pos: positive; Neg: negative.; *Ct-value cut-off at which p > 0.5 for a positive viral culture result.

|  | Sensitivity (95%CI) |  |  | Specificity (95%CI) |  |  |
| --- | --- | --- | --- | --- | --- | --- |
|  | BD | Roche | Overall | BD | Roche | Overall |
| qRT-PCR pos. or neg. | 48.9% (41.6-56.2) | 61.5% (54.4-68.1) | 55.4% (50.3-60.4) | 99.9% (99.5-100) | 99.7% (99.3-99.9) | 99.8% (99.5-99.9) |
| qRT-PCR, Ct - value < cutoff* | 75.5% (66.6-82.6) | 80.1% (72.7-86.0) | 78.0% (72.5-82.8) | 99.7% (99.2-99.8) | 99.1% (98.5-99.5) | 99.4% (99.0-99.6) |
| Composite reference standard | 75.0% (64.7-83.1) | 81.4% (74.2-86.9) | 76.8% (70.4-82.2) | 99.8% (99.5-100.0) | 99.7% (99.3-99.9) | 99.8% (99.5-99.9) |

**Table 3.** Uni-and multivariate logistic regression analysis for the occurrence of false negative self-test compared to RT-PCR result; ^⍰^ (Yes: 1; No: 0);⍰ (1 / 2: 1; 3 / 4: 0).

| Variable | Univariate analysis |  | Multivariate analysis |  |
| --- | --- | --- | --- | --- |
|  | Odds ratio (95%CI) | p-value | Odds ratio (95%CI) | p-value |
| Ct-value lower than Ct-value cut-off▢ | 0.032 (0.017-0.061) | <0.001 | 0.032 (0.016-0.063) | <0.001 |
| Roche RDT▢ | 0.637 (0.423-0.960) | 0.031 | 0.665 (0.384-1.152) | 0.146 |
| Current COVID-19 related symptoms▢ | 0.401 (0.246-0.652) | <0.001 | 0.696 (0.356-1.361) | 0.290 |
| Male▢ | 0.654 (0.434-0.986) | 0.043 | 0.696 (0.356-1.093) | 0.100 |
| Age (Years) | 1.012 (0.999-1.0) | 0.076 | 1.021 (1.002-1.040) | 0.026 |
| Ease (0 (very difficult)- 10 (very easy)) | 0.908 (0.802-1.029) | 0.131 | 0.843 (0.712-1.000) | 0.049 |
| Confidence in result interpretation▢ | 7.191 (0.912-57.344) | 0.063 | 6.439 (0.503-82.4734) | 0.152 |
| Recommend self-testing▢ | 0.895 (0.295-2.715) | 0.844 |  |  |
| Education level (1-4) | 1.158 (0.862-1.556) | 0.329 |  |  |
| Watched the instruction video▢ | 0.983 (0.634-1.526) | 0.941 |  |  |
| The instructions for use were clear▢ | 1.545 (0.139-17.187) | 0.723 |  |  |
| confident in performing the nose sampling properly▢ | 0.757 (0.320-1.793) | 0.527 |  |  |
| confident in performing the RDT properly▢ | 1.824 (0.464-7.163) | 0.389 |  |  |
| Would use this rapid test again▢ | 0.895 (0.295-2.715) | 0.984 |  |  |

## Discussion

Self-testing using commercially available antigen tests proved to be feasible and delivered reliable results. Specificity was extremely high (>99%) while sensitivities were 75.5% (BD-RDT) and 80.1%. (Roche-RDT) respectively. In addition, we identified determinants of false-negative results which may offer opportunities for further improvement of performance. For example, the sensitivity was higher in younger participants and also in participants whom found the self-testing easy. Future studies could target specific age groups and see if this improves performance. Participants also frequently indicated that they were uncertain if they had inserted the swab far enough into the nose. We used a standard nasal swab, an adapted design of the swab that indicates the 2,5 cm insertion on the swab could potentially further improve the quality of sampling.

To assess the performance of the RDT’s we determined the CT-value at which viable virus could readily be detected. The two PCR platforms showed similar CT-value patterns for samples with positive and negative viral cultures with minor differences in the threshold for positive viral culture. We used viral culture as a reference to determine the cut off as we aimed to determine the sensitivity of the RDT’s to detect infectious individuals. PCR is a highly sensitive method that can detect viral RNA for prolonged periods after the initial infection. Most of these individuals however, are no longer infectious (15). There is an ongoing discussion about the value of PCR with high CT values for this kind of evaluations as it is clear that most high CT values do not represent infectious cases. However, the exact threshold is unclear and varies between PCR platforms (19).For the two PCR platforms used in this study the thresholds for the presence of viable virus were determined at 23.0 for the AA and 24.5 for the LDA. These values cannot be extrapolated to other platforms.

There are limited published data on the reliability of self-testing. A recent study conducted in Germany, found that a layperson can be trained to administer a rapid self-test properly (11). The study involved 146 individuals showing symptoms of which 40 tested positive for SARS-CoV-2 using a PCR test. All subjects then conducted additional self-tests using nasal swabs. Of all those who tested positive, 91,4% were able to confirm their result via rapid self-test. And practically all of those who tested negative were able to confirm their result with a self-test. This shows the potential of self-testing. However, it was not a real-life evaluation. As far as we are aware, we present the first large-scale clinical evaluation of self-testing for SARS-CoV-2. The results show that self-testing is possible and delivers useful results. Not for medical diagnostic testing as the sensitivity is not optimal. However, the test characteristics are suitable for large scale preventive testing programs to open up specific activities in society. A modelling study showed that for preventive testing the frequency of testing is much more important than the sensitivity of the test (5). For example, a test with 80% sensitivity performed by at least 70% of the population once weekly was estimated to reduce the Rt from 1.5 to below 1.0. This may facilitate opening up activities like education, contact professions, high-risk jobs (e.g., slaughterhouses) and so on. These theoretical considerations should be confirmed in real-life settings.

In conclusion, we showed that self-testing using currently available RDT’s has a high specificity and relatively high sensitivity (75%-80%) to identify individuals with a high probability of contagiousness. This application has the potential for frequent and extensive testing which may be an aid to lift current restrictions to society.

## Data Availability

All data has been included in the manuscript

## Acknowledgements

We would like to thank all health-care, call-centre and supporting staff of the GGD test centre in Tilburg, the Netherlands for their help in retrieving the samples and completion of the surveys. We would like to thank JvK and BP for their help in coordinating and retrieving the informed consent, telephone numbers and e-mail addresses of the participants. Additionally, we would like to thank the laboratory technicians of Microvida for their help in performing the RT-PCR’s.

## References

1. Zhu N, Zhang D, Wang W, Li X, Yang B, Song J, et al. A Novel Coronavirus from Patients with Pneumonia in China, 2019. N Engl J Med [Internet]. 2020 Feb 20;382(8):727–33. Available from: http://www.nejm.org/doi/10.1056/NEJMoa2001017

2. Huang C, Wang Y, Li X, Ren L, Zhao J, Hu Y, et al. Clinical features of patients infected with 2019 novel coronavirus in Wuhan, China. Lancet [Internet]. 2020 Feb;395(10223):497–506. Available from: https://linkinghub.elsevier.com/retrieve/pii/S0140673620301835

3. Grassly NC, Pons-Salort M, Parker EPK, White PJ, Ferguson NM, Ainslie K, et al. Comparison of molecular testing strategies for COVID-19 control: a mathematical modelling study. Lancet Infect Dis [Internet]. 2020 Dec;20(12):1381–9. Available from: https://linkinghub.elsevier.com/retrieve/pii/S1473309920306307

4. Paltiel AD, Zheng A, Walensky RP. Assessment of SARS-CoV-2 Screening Strategies to Permit the Safe Reopening of College Campuses in the United States. JAMA Netw open. 2020;3(7):e2016818.

5. Bootsma M, Kretzschmar M, Rozhnova G, Heesterbeek J, Kluytmans J, Bonten M. Regular universal screening for SARS-CoV-2 infection may not allow reopening of society after controlling a pandemic wave. medRxiv. 2020;

6. Larremore DB, Wilder B, Lester E, Shehata S, Burke JM, Hay JA, et al. Test sensitivity is secondary to frequency and turnaround time for COVID-19 screening. Sci Adv [Internet]. 2021 Jan;7(1):eabd5393. Available from: https://advances.sciencemag.org/lookup/doi/10.1126/sciadv.abd5393

7. Van der Moeren N, Zwart VF, Lodder EB, Bijllaardt W Van den, Esch HRJM Van, Stohr JJJM, et al. Performance evaluation of a SARS-CoV-2 rapid antigentest: Test perfomance in the community in the Netherlands. medRxiv. 2020;1–13.

8. Gremmels H, Winkel BMF, Schuurman R, Rosingh A, Rigter NAM, Rodriguez O, et al. Real-life validation of the PanbioTM COVID-19 antigen rapid test (Abbott) in community-dwelling subjects with symptoms of potential SARS-CoV-2 infection. EClinicalMedicine [Internet]. 2021 Jan;31:100677. Available from: https://doi.org/10.1016/j.eclinm.2020.100677

9. Igloi Z, Velzing J, van Beek J, van de Vijver D, Aron G, Ensing R, et al. Clinical evaluation of the Roche/SD Biosensor rapid antigen test with symptomatic, non-hospitalized patients in a municipal health service drive-through testing site. medRxiv. 2020;1–15.

10. Lindner AK, Nikolai O, Rohardt C, Burock S, Hülso C, Bölke A, et al. Head-to-head comparison of SARS-CoV-2 antigen-detecting rapid test with professional-collected anterior nasal versus nasopharyngeal swab. medRxiv. 2020;

11. Lindner AK, Nikolai O, Rohardt C, Kausch F, Wintel M, Gertler M, et al. SARS-CoV-2 patient self-testing with an antigen-detecting rapid test: a head-to-head comparison with professional testing. medRxiv [Internet]. 2021;2021.01.06.20249009. Available from: https://doi.org/10.1101/2021.01.06.20249009

12. Hoehl S, Schenk B, Rudych O, Göttig S, Foppa I, Kohmer N, et al. At-home self-testing of teachers with a SARS-CoV-2 rapid antigen test to reduce potential transmissions in schools Results of the SAFE School Hesse Study. medRxiv. 2020;

13. Figueroa C, Johnson C, Ford N, Sands A, Dalal S, Meurant R, et al. Reliability of HIV rapid diagnostic tests for self-testing compared with testing by health-care workers: a systematic review and meta-analysis. Lancet HIV [Internet]. 2018;5(6):e277–90. Available from: http://dx.doi.org/10.1016/S2352-3018(18)30044-4

14. Wölfel R, Corman VM, Guggemos W, Seilmaier M, Zange S, Müller MA, et al. Virological assessment of hospitalized patients with COVID-2019. Nature. 2020;581(7809):465–9.

15. van Kampen JJA, van de Vijver DAMC, Fraaij PLA, Haagmans BL, Lamers MM, Okba N, et al. Duration and key determinants of infectious virus shedding in hospitalized patients with coronavirus disease-2019 (COVID-19). Nat Commun [Internet]. 2021;12(1):8–13. Available from: http://dx.doi.org/10.1038/s41467-020-20568-4

16. Sikkema RS, Pas SD, Nieuwenhuijse DF, O’Toole Á, Verweij JJ, van der Linden A, et al. COVID-19 in health-care workers in three hospitals in the south of the Netherlands: a cross-sectional study. Lancet Infect Dis. 2020;20(11):1273–80.

17. Corman VM, Landt O, Kaiser M, Molenkamp R, Meijer A, Chu DKW, et al. Detection of 2019 novel coronavirus (2019-nCoV) by real-time RT-PCR. Eurosurveillance. 2020;25(3):1–8.

18. Scheltinga SA, Templeton KE, Beersma MFC, Claas ECJ. Diagnosis of human metapneumovirus and rhinovirus in patients with respiratory tract infections by an internally controlled multiplex real-time RNA PCR. J Clin Virol [Internet]. 2005 Aug;33(4):306–11. Available from: https://linkinghub.elsevier.com/retrieve/pii/S1386653205001393

19. Guglielmi G. Rapid coronavirus tests: a guide for the perplexed. Nature [Internet]. 2021 Feb 11;590(7845):202–5. Available from: http://www.nature.com/articles/d41586-021-00332-4

