## Supplementary methods S1 for "Self-testing for the detection of SARS-CoV-2 infection with rapid antigen tests"

### COVID-19
### HOMETEST

#### INSTRUCTIONS

**Perform the test immediately after arrival at home**

**Questions or help needed?**
Call (0)85 0659916
www.corona-test-instructies.nl

GGD

#### Thank you for your cooperation!

This study investigates whether people are able to perform a COVID-19 rapid test themselves at home and whether it gives a reliable result.

The results of the research are important to society, so we greatly appreciate your cooperation. Read the instructions carefully before you start the test.

Watch the short instruction video at: **www.corona-test-instructies.nl**

**You will receive an email from the sender "Castor EDC" with subject "Home Test Examination Questionnaire" sent to your specified e-mail address.**

Please complete this questionnaire immediately after you have obtained the result of the test. If you have any questions, you can contact the home test information number at (0) 85 0659916

Sincerely,
Prof. Dr. J.A.J.W. Kluytmans, project lead

**WATCH THE VIDEO**

#### Your test kit consists of:

##### Test device

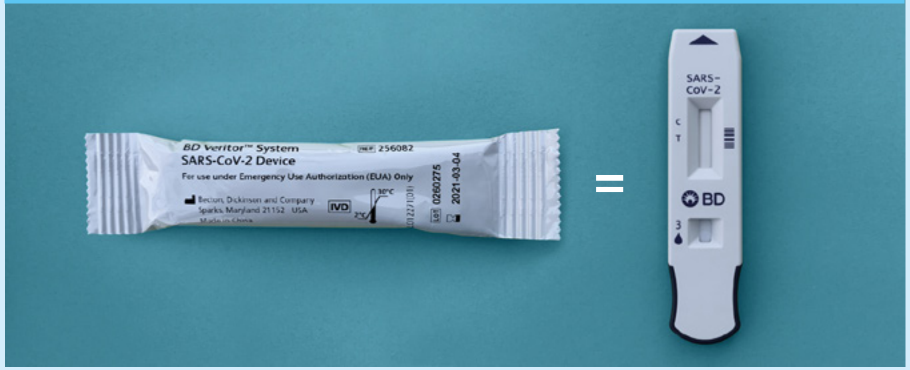


##### Sterile swab

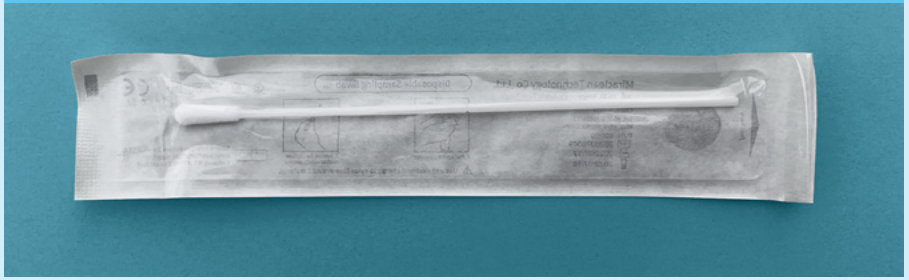

##### Buffer tube

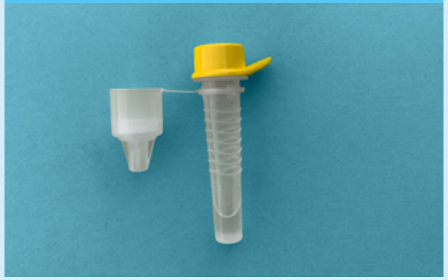

##### Cardboard tube holder

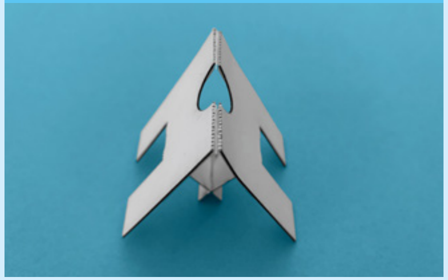

### COVID-19 **HOME TEST**

#### PREPARATIONS

Watch the video: **www.corona-test-instructies.nl**

##### 1. HYGIENE

- Blow your nose
- Wash your hands

##### 2. TUBE HOLDER

- Fold the cardboard holder together, as shown below

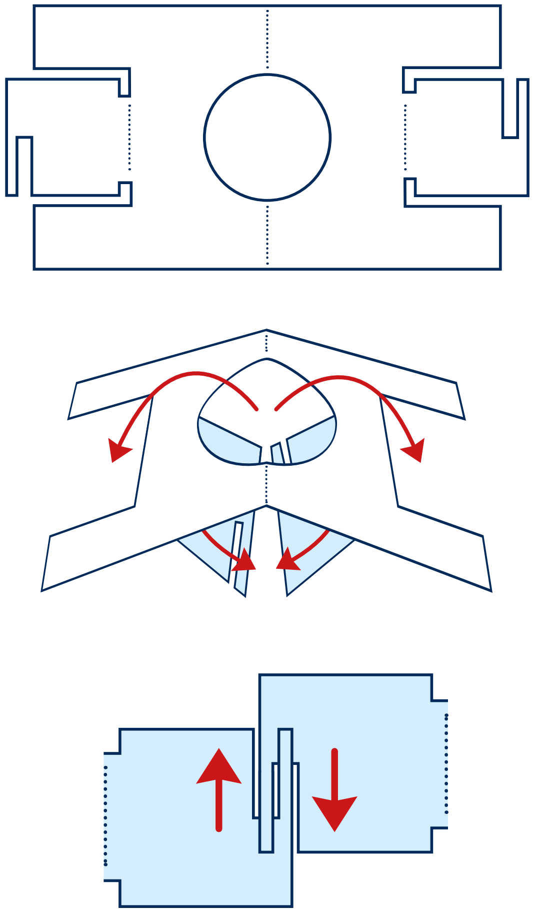

##### 3. SET UP

- Remove the rapid test from the packaging
- Remove the cotton swab from the packaging
- Carefully remove the cellophane from the liquid tube
- Place the tube in the holder

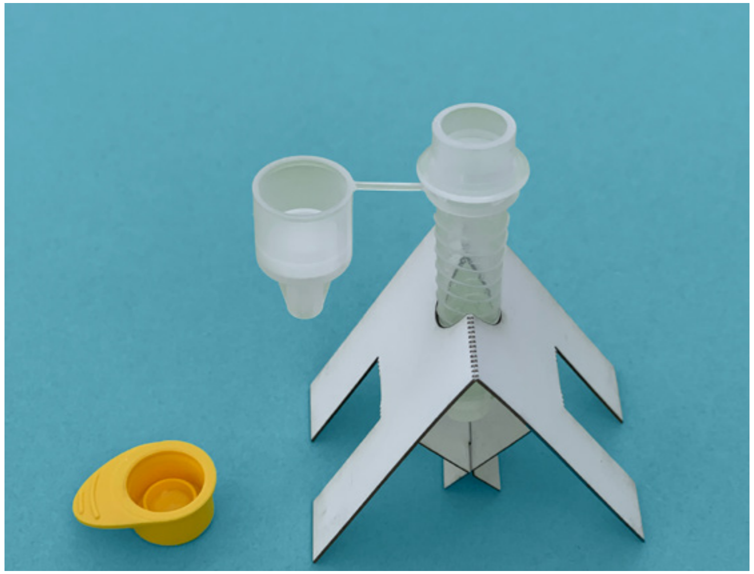

If opening the rapid test or cotton swab is challenging, use a pair of scissors.

### COVID-19 **HOME TEST**

#### TEST PROCEDURE

##### 4. MEASURE

- Hold the cotton swab at 2.5 cm from the top (including the tip of the swab)
- Use the ruler below

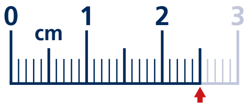


##### 5. NOSE SWAB

- Put the cotton swab gently **about 2.5 cm** into the nostril
- Rotate the swab 5 times against the nasal wall, rubbing with some resistance
- Your eyes might tear up, but this is normal

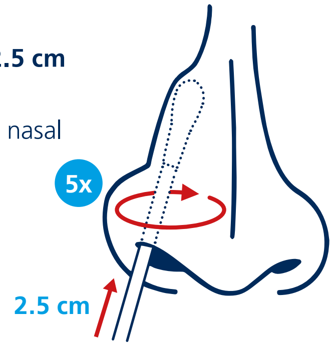


##### 6. OTHER NOSTRIL

- Put the same end of the same swab gently **about 2.5 cm** into the other nostril
- Rotate the swab 5 times against the nasal wall, rubbing with some resistance
- Your eyes might tear up, but this is normal

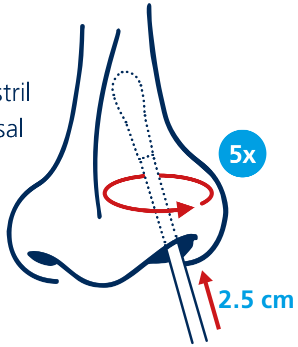


##### 7. MIX CAREFULLY

DON'T SPILL!

- Insert the swab into the liquid of the buffer tube
- Gently move the swab up and down for about 15 seconds

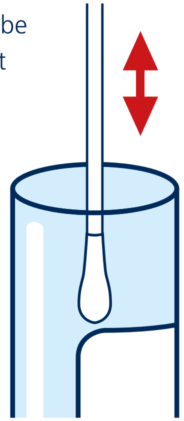

##### 8. NOZZLE CAP

- Press the nozzle cap tightly onto the tube till you hear a **'click'**.

##### 9. DROPS

- Apply 3 drops of the extracted sample in the small well of the test device
- Set a timer for 15 minutes

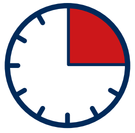


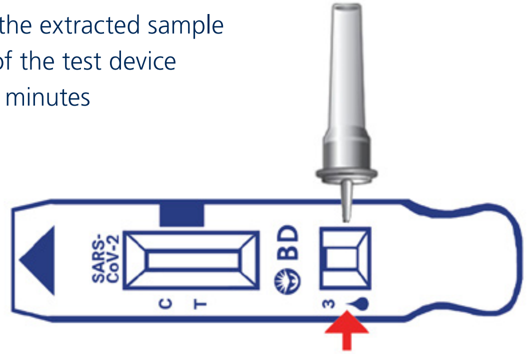

### COVID-19 **HOME TEST**

#### INTERPRETING RESULTS

##### 10. RESULTS

- Read the test result at 15 minutes after you have added the drops to the cartridge
- Warning: risk of incorrect results. Do not read the test result after 30 min.

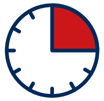

**15 minutes**

###### POSITIVE

- If 2 colored lines appear, the result is **POSITIVE**
- You are **most likely** infected with Covid-19
- Stay at home and wait for the results of the test lane to confirm your infection

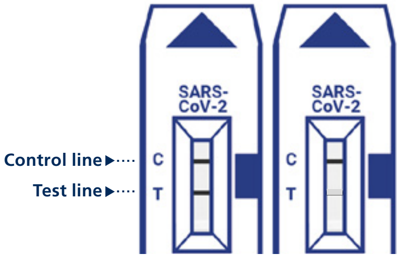


###### NEGATIVE

- If you see 1 colored line in the top-section the result is **NEGATIVE**
- You are **most likely** not infected with Covid-19
- Stay at home and wait for the results of the test lane to make sure you are not infected

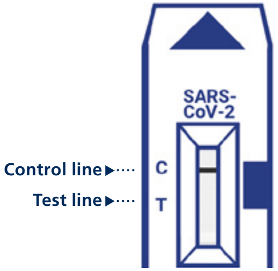


###### INVALID

- If you see 1 colored line at the bottom section the result is **INVALID**
- Stay inside and wait for the results of the test lane

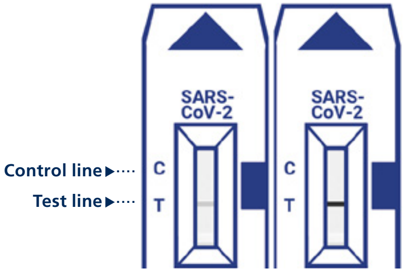


##### 11. FILL OUT THE ONLINE FORM

- You will receive an email from the sender "Castor EDC" with subject "Home Test Examination Questionnaire" sent to your specified e-mail address.
- This email contains a link to the online questionnaire
- Fill out the questionnaire right after you performed the test

##### 12. QUESTIONS?

- If you have any questions, you can contact the home test information number at (0) 85 0659916

##### 13. GARBAGE

- You can put the materials in your own waste bin

#### Frequently Asked Questions

**What does the "C" on the test mean?**
That is the c of control line, this is an indicator to show there is enough liquid in the test.

**More than 3 drops came out, now what?**
That is okay, if the test "fills up" (light pink) and if the colored line at the C is shown, it is good.

**Foam came out with the drops, now what?**
That is okay, if the test "fills up" (light pink) and there is a line on the C, it is good.

**After a few hours my test was different**
The result after 15 minutes (and no later than 30 minutes) is the result that applies.

**My test lane result was different from the rapid test, now what?**
The result of the test lane (PCR) is leading. Follow the GGD instructions.

**Other questions?**
Call the home test information number: (0) 85 0659916

### COVID-19 **HOME TEST**

#### GENERAL INFORMATION

##### 1. General Information
This study is a collaboration between the participating hospitals Breda (Amphia) / Tilburg (Elisabeth-TweeSteden hospital), the Ministry of Health, Welfare and Sport, the municipal health services (GGD) and the National Institute for Public Health and the Environment (RIVM)

##### 2. The Purpose of the Study
Investigate the clinical performance of the antigen rapid tests when used for self-testing at home and to determine whether individuals correctly interpret the test result themselves.

##### 3. Background
Currently, so-called PCR tests are used for people with symptoms of coronavirus disease (COVID-19). A PCR test is sensitive and precise but analyzing is only possible in specialized laboratories. People need to wait relatively long for the result and currently, laboratories are under pressure.

Last October, a rapid antigen test was validated in the test lanes of GGD Breda, in collaboration with the Amphia hospital and the Ministry of Health, Welfare and Sport. This antigen test has a good sensitivity and gives a result in 15 minutes. Since then, antigen rapid tests have been in great demand.

In this study, we investigate if rapid antigen tests can be processed and read by people with symptoms of COVID-19. This could save a lot of time. In addition, we will check whether the test results correspond with the results of the conducted rapid test study and relate to the result of the PCR test.

##### 4. What happens when you participate?
If you develop symptoms that could indicate COVID-19, you make an appointment to get tested. When you arrive at the test lane, you may be asked if you want to take part in this study. If you decide to participate, you will receive a PCR test (hereafter referred to as: "test lane test"). In addition, you will receive a package containing the test kit and instructions how to take the sample, process the test and then interpret the results.

You are supposed to process the rapid antigen test when you arrive at home. After taking the sample, and processing the sample with the test kit, you interpret the results after 15 minutes. You fill out a few questions and the test results through an online questionnaire.

If you have a POSITIVE test result, you are probably infected with COVID-19. In that case, you must stay home, remain in home quarantine, and wait for the results of the test lane test.

If you have a NEGATIVE test result, you are probably not infected with COVID-19. But you must stay home, remain in home quarantine, and wait for the results of the test lane test.

The result of the test lane is leading. The test result of the test lane will follow within 48 hours. In some cases, the test results may differ from the results of the rapid antigen test.

##### 5. Potential risks and side effects
Collecting a nasal swab sample might feel uncomfortable and eyes could tear up for a little while. Apart from a time investment of 20 minutes, participation does not have any disadvantages.

There are no direct personal benefits related to your participation. Though you have a faster confirmation when you are infected with the COVID-19 virus. Information obtained from this study will help to improve the COVID-19 test strategy. You will not be reimbursed or paid for participation in this study.

##### 6. If you do not wish to participate or would like to stop
Participation is voluntary. If you participate, you can always change your mind and still stop, even if you already agreed. You do not have to motivate why you wish to cancel. Please inform the researcher when you decide to stop. The data collected up to that point will be used for the research.

##### 7. Study Sample Storage and Retention
For this study, your personal data and bodily material (the sample taken on the test lane) will be processed and stored. This includes personal data such as your name, age and the information stated under point 4. The use and storage of your data and your bodily material is necessary to answer the questions addressed in this study and being able to publish the (non-traceable) results. This data will only be used for this purpose and not be shared with others. We ask your permission for the use of your data and bodily material.

##### 8. Insurance policy
There is no extra risk involved for participants of the study.

##### 9. Questions?
If you have any questions, please contact the home test information number:
0) 85 0659916. For independent advice on participating in this study, please contact Dr. A.J.P. Joosten, on the number 076 - 5954192.

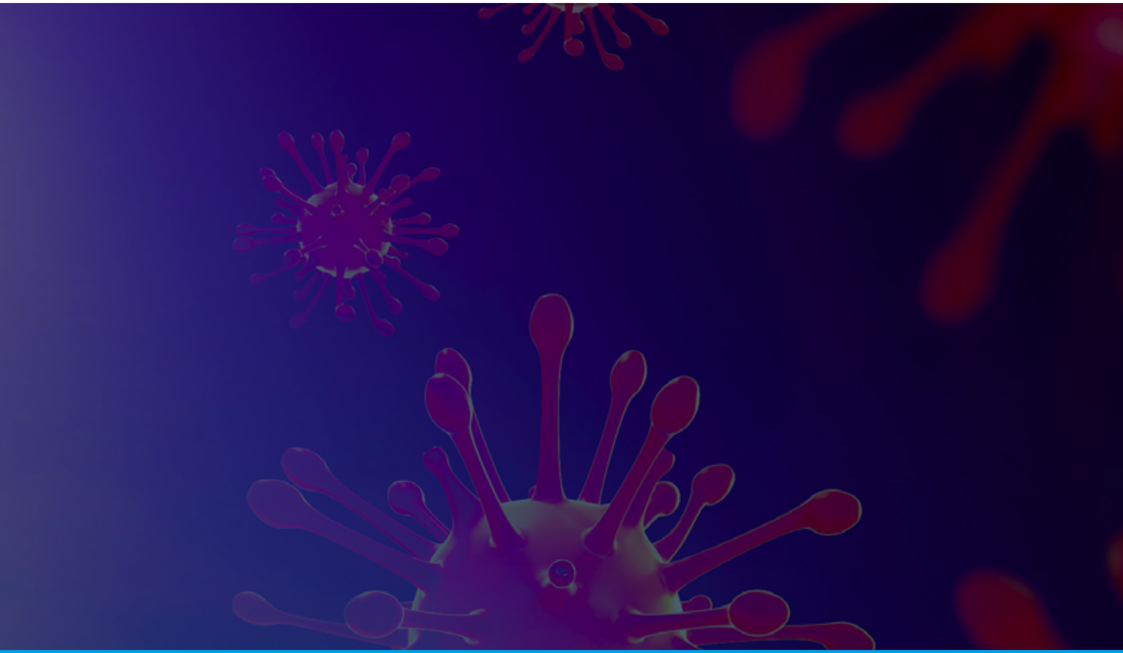

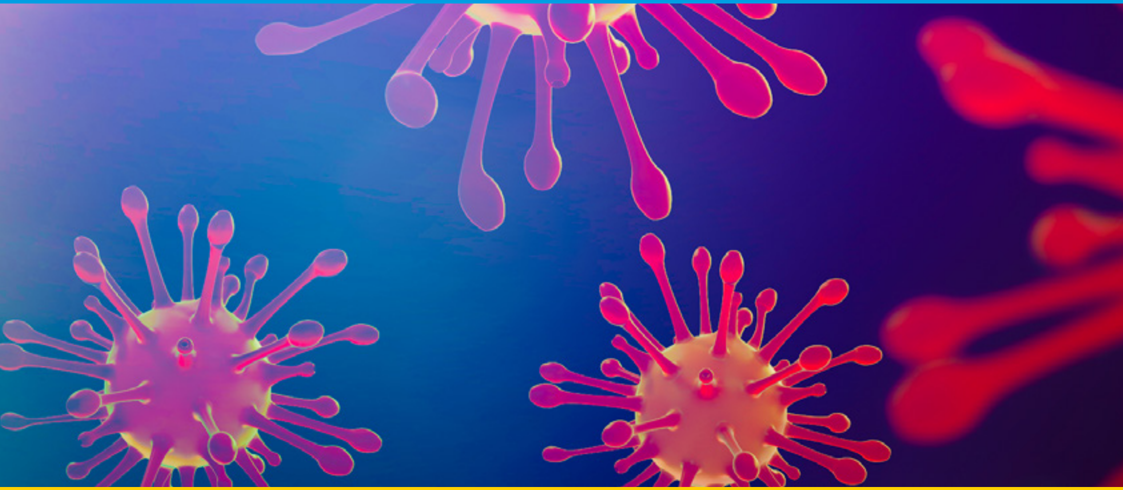

MICROVIDA
MEDISCHE MICROBIOLOGIE BRABANT & ZEELAND

Ministerie van Volksgezondheid, Welzijn en Sport

GGD
