## Supplementary methods S2 for "Self-testing for the detection of SARS-CoV-2 infection with rapid antigen tests"

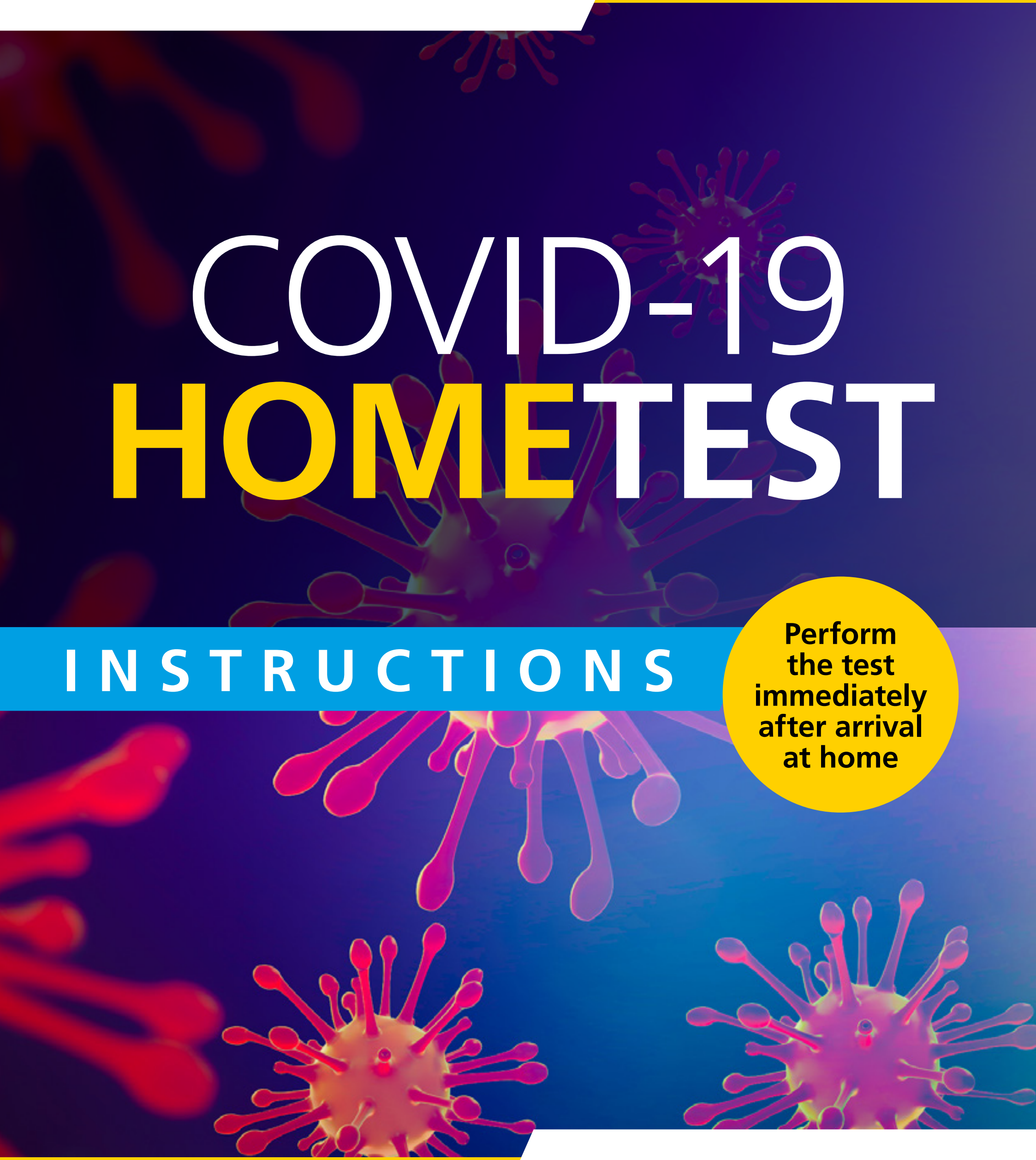

### COVID-19 HOMETEST

#### INSTRUCTIONS

**Perform the test immediately after arrival at home**

**Questions or help needed?**

**Call (0)85 0659916**
**www.coronatest-instructies.nl**

GGD

### COVID-19 **HOME TEST**

INSTRUCTIONS

#### Thank you for your cooperation!

This study investigates whether people are able to perform a COVID-19 rapid test themselves at home and whether it gives a reliable result.

The results of the research are important to society, so we greatly appreciate your cooperation. Read the instructions carefully before you start the test.

Watch the short instruction video at: **www.coronatest-instructies.nl**

**You will receive an email from the sender “Castor EDC” with subject “Home Test Examination Questionnaire” sent to your specified e-mail address.**

Please complete this questionnaire immediately after you have obtained the result of the test. If you have any questions, you can contact the home test information number at (0) 85 0659916

Sincerely,
Prof. Dr. J.A.J.W. Kluytmans, project lead

**WATCH THE VIDEO**

#### Your test kit consists of:

##### Test device

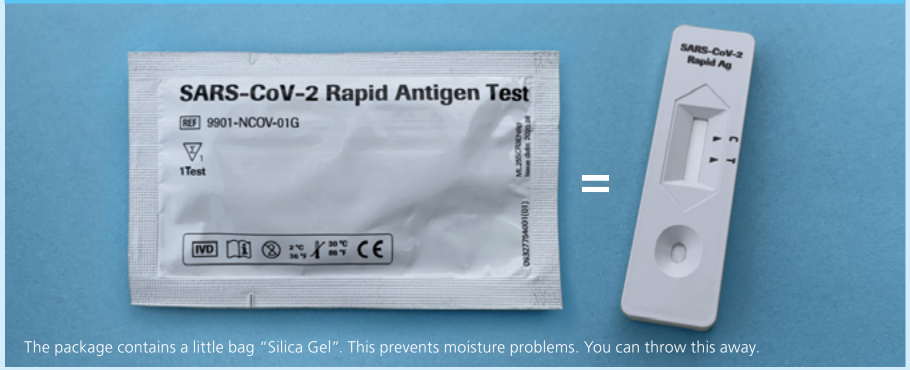


The package contains a little bag “Silica Gel”. This prevents moisture problems. You can throw this away.

##### Sterile swab

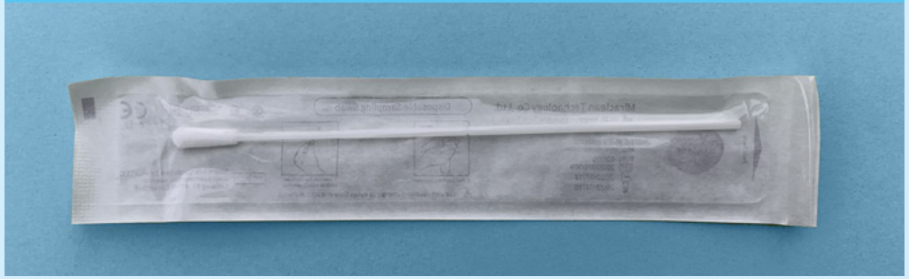

##### Buffer tube

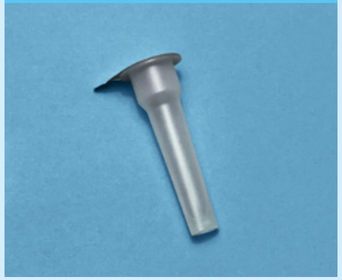

##### Nozzle cap

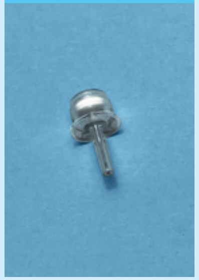

##### Cardboard tube holder

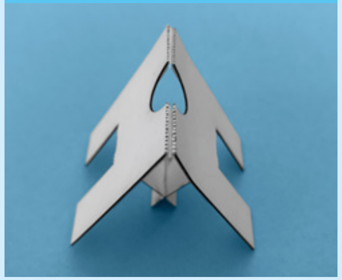

### COVID-19 **HOME TEST**

#### PREPARATIONS

Watch the video: **www.coronatest-instructies.nl**

#### 1. HYGIENE

- Blow your nose
- Wash your hands

#### 2. TUBE HOLDER

- Fold the cardboard holder together, as shown below

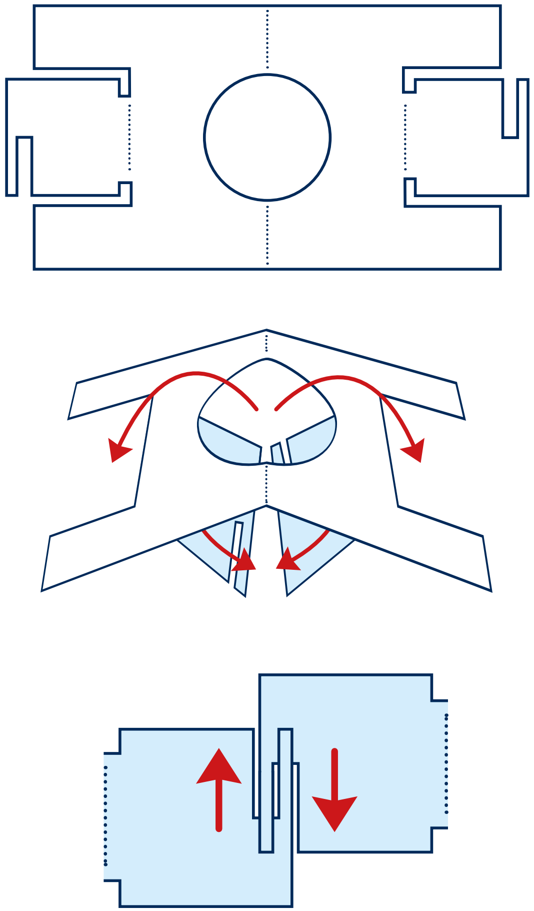

#### 3. SET UP

- Remove the rapid test from the packaging
- Remove the cotton swab from the packaging
- Carefully remove the cellophane from the liquid tube
- Place the tube in the holder

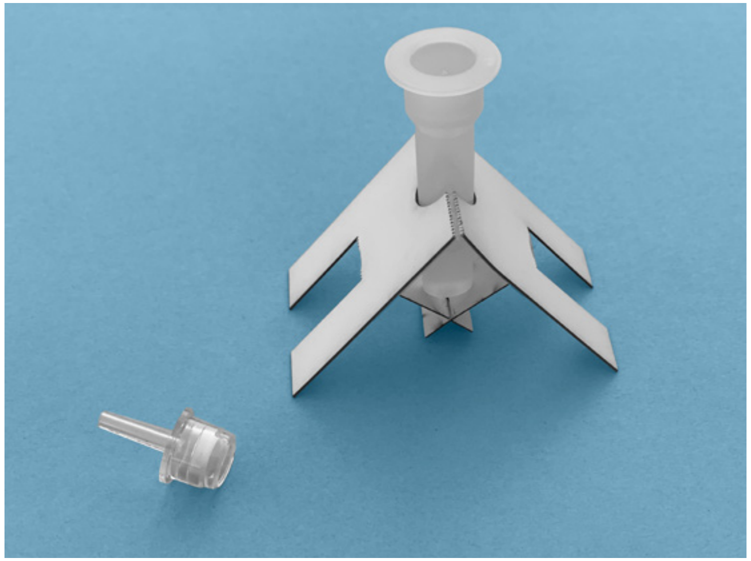

If opening the rapid test or cotton swab is challenging, use a pair of scissors.

### COVID-19 **HOME TEST**

#### TEST PROCEDURE

##### 4. MEASURE

- Hold the cotton swab at 2.5 cm from the top (including the tip of the swab)
- Use the ruler below

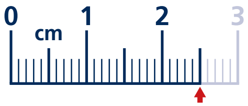


##### 5. NOSE SWAB

- Put the cotton swab gently **about 2.5 cm** into the nostril
- Rotate the swab 5 times against the nasal wall, rubbing with some resistance
- Your eyes might tear up, but this is normal

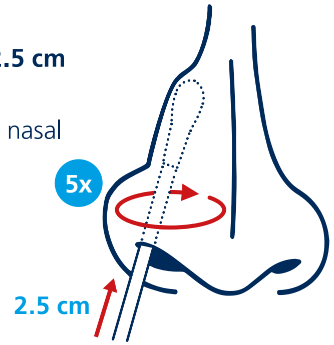


##### 6. OTHER NOSTRIL

- Put the same end of the same swab gently **about 2.5 cm** into the other nostril
- Rotate the swab 5 times against the nasal wall, rubbing with some resistance
- Your eyes might tear up, but this is normal

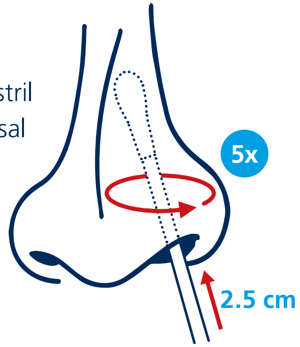


##### 7. MIX CAREFULLY

- Insert the swab into the buffer tube
- While squeezing the buffer tube, stir the swab more than 5 times.
- Remove the swab while squeezing the sides of the tube to extract **all the liquid** from the swab

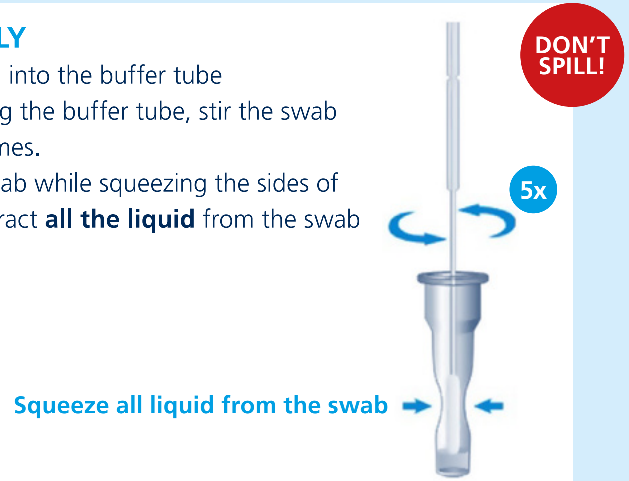


##### 8. NOZZLE CAP

- Press the nozzle cap tightly onto the tube.

##### 9. DROPS

- Apply 3 drops of the extracted sample in the small well of the test device
- Set a timer for 15 minutes

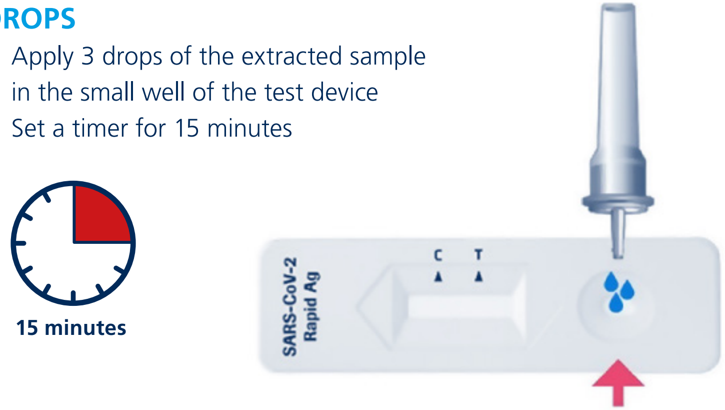

### COVID-19 **HOME TEST**

#### INTERPRETING RESULTS

##### 10. RESULTS

- Read the test result at 15 minutes after you have added the drops to the cartridge
- Warning: risk of incorrect results. Do not read the test result after 30 min.

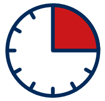

**15 minutes**

###### POSITIVE

- If 2 colored lines appear, the result is **POSITIVE**
- You are **most likely** infected with Covid-19
- Stay at home and wait for the results of the test lane to confirm your infection

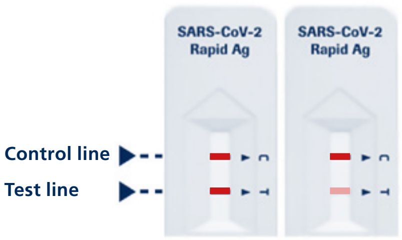


###### NEGATIVE

- If you see 1 colored line in the top-section the result is **NEGATIVE**
- You are **most likely** not infected with Covid-19
- Stay at home and wait for the results of the test lane to make sure you are not infected

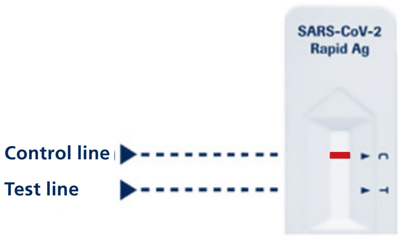


###### INVALID

- If you see 1 colored line at the bottom section the result is **INVALID**
- Stay inside and wait for the results of the test lane

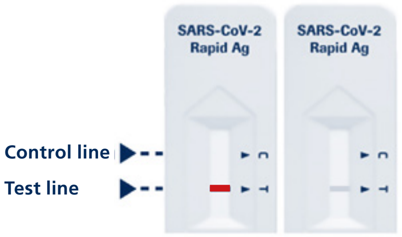


##### 11. FILL OUT THE ONLINE FORM

- You will receive an email from the sender "Castor EDC" with subject "Home Test Examination Questionnaire" sent to your specified e-mail address.
- This email contains a link to the online questionnaire
- Fill out the questionnaire right after you performed the test

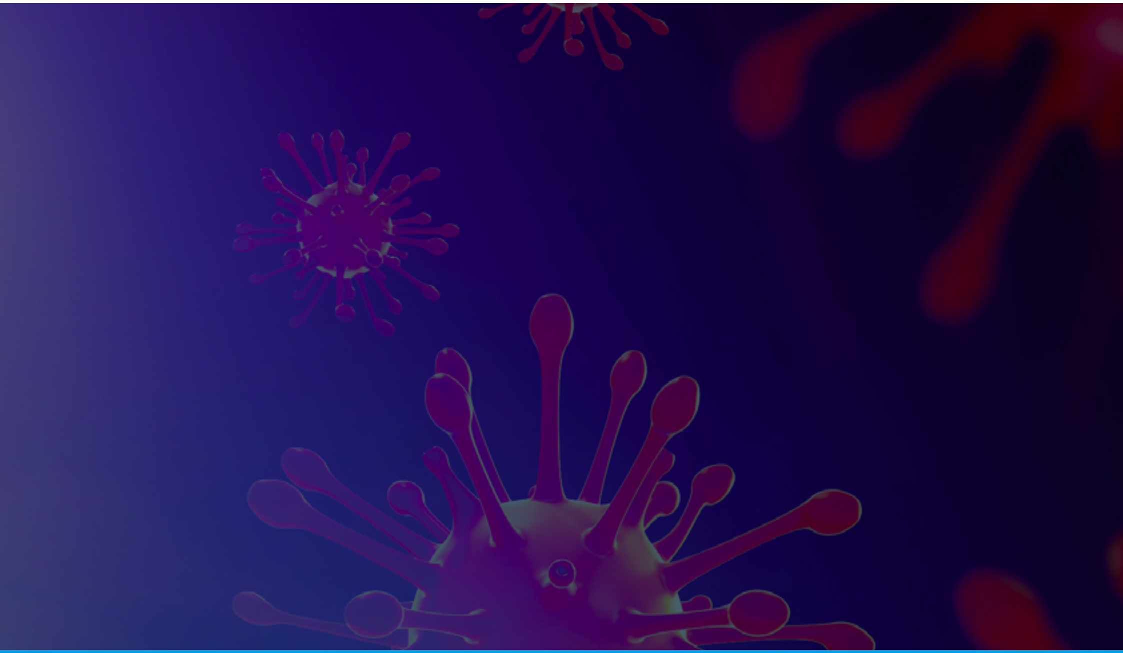

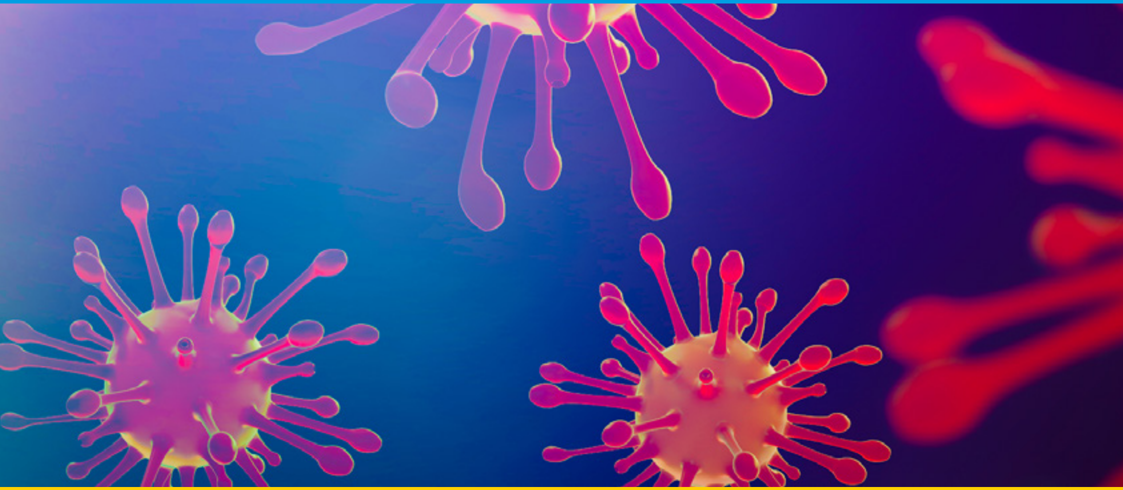

MICROVIDA
MEDISCHE MICROBIOLOGIE BRABANT & ZEELAND

Ministerie van Volksgezondheid,
Welzijn en Sport

GGD
