## Supplementary methods S3 for "Self-testing for the detection of SARS-CoV-2 infection with rapid antigen tests"

Survey:

Test result and general questions

1. What is the result of the rapid antigen test? (positive / negative)
2. Have you experienced COVID-19 related symptoms in the past 3 weeks? (yes / no)
3. Do you currently experience COVID-19 related symptoms? (yes / no)
4. What is your age (age in years)?
5. What is your sex (male/female / other)?
6. What is your highest level of education (elementary school / high school / bachelor degree / master degree or higher)?

Questions regarding performing the test:

1. Have you watched the instruction video? (yes / no)
2. On a scale from 0 to 10: how difficult did you find the procedure of the test (0 to 10)

Statements,

Answer options:

- Totally agree
- Partially agree
- Partially disagree
- Totally disagree

1. The instructions for use were clear
2. I am confident I performed the nose sampling properly
3. I am confident I performed the execution of test properly
4. I am confident I interpreted the test result properly
5. When again experiencing COVID-19 related symptoms I would use this rapid test again
6. I would recommend this rapid test to other persons experiencing COVID-19 related symptoms
