## Supplementary figures and images for "Self-testing for the detection of SARS-CoV-2 infection with rapid antigen tests"

### Supplementary figure S1

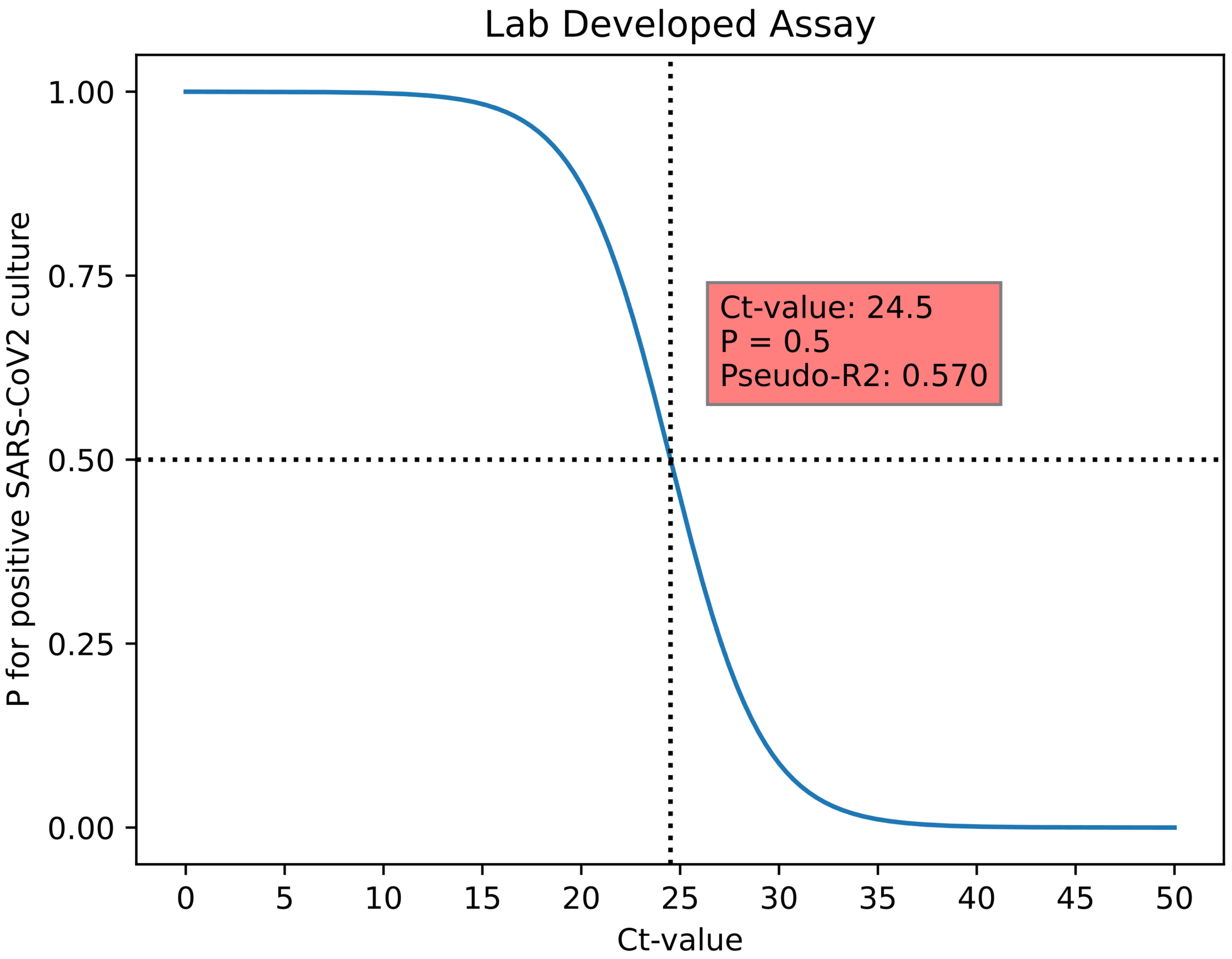

### Supplementary figure S2

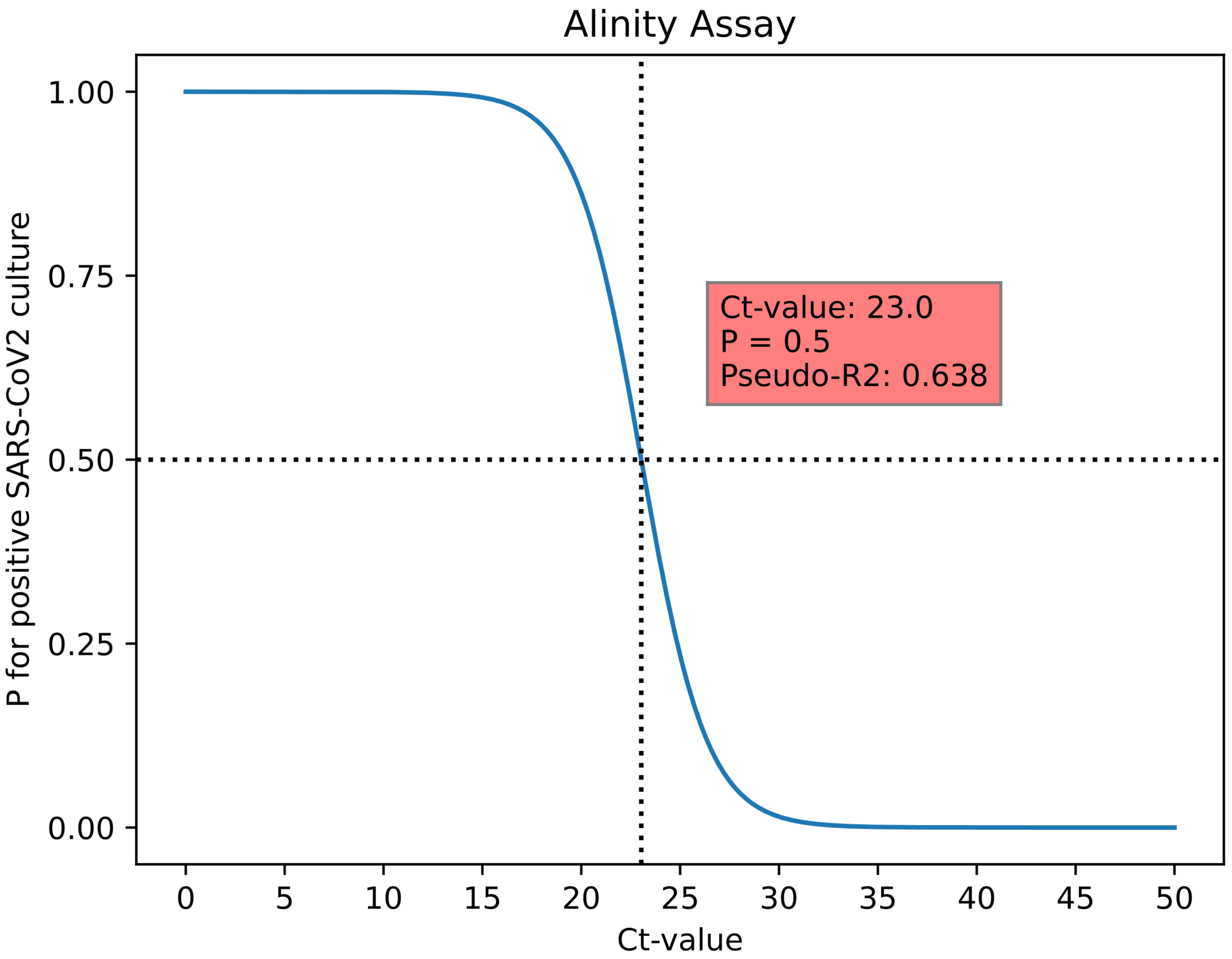
